# Predictors and outcomes of pulmonary hypertension after mitral valve replacement in severe mitral regurgitation

**DOI:** 10.1101/2024.10.07.24315064

**Authors:** García-Orta Rocío, González Molina Mercedes, Uribe Morales Inés, Torres Quintero Lucía, Rodríguez Torres Diego J., Segura Rodríguez Diego, Moreno Escobar Eduardo

## Abstract

**Background:** The objective of the study was to determine perioperative clinical and echocardiographic factors associated with pulmonary hypertension (PH) during long-term follow-up of patients with severe primary mitral regurgitation (MR) after mitral valve replacement surgery and its prognostic repercussions.

**Methods:** Prospective cohort of consecutive patients undergoing mitral valve replacement between January 2005 and January 2015 and followed for ≥3 years. Main exclusion criteria were receipt of mitral repair surgery and the presence of mitral stenosis. The main outcome was a systolic pulmonary pressure (SPAP) by echocardiography >50 mmHg at least 12 months’ post-surgery.

**Results:** 106 patients, 46% male, mean age of 61.7±11 yrs. and median follow-up of 8.8 [6- 16] yrs. SPAP was >50 mmHg in 33% of patients. Predictors identified by multivariate analysis were diabetes (OR 8.3, 95%CI 1.1-73.9), persistent atrial fibrillation (OR 4.2, 95%CI 1.1-20.6), non-rheumatic MR (OR 7.3, 95%CI 1.1-45.5), prosthesis gradient, (OR 2.8, 95%CI 1.3-5.7), patient-prosthesis mismatch (OR 16.9, 95%CI 1.2-234), and preoperative SPAP ≥50 mmHg (OR 19.3, 95%CI 3.4-107). Patients with PH had higher hospitalization (78.8 % vs. 37%, p<0.001) and mortality (72,4% vs. 19,7%, p<0.001) rates. The 10-year survival rate was 83% in patients without PH *versus* 30% in those with PH (p<0.001).

**Conclusions:** Persistent PH is a frequent and serious long-term complication of valve replacement for severe mitral regurgitation. It is associated with a poor prognosis and is related to preoperative pulmonary pressure. Surgery could be considered in candidates for mitral valve replacement with moderate PH, especially in the presence of atrial fibrillation.

**Clinical perspective:** *What is already known on this topic:* Pulmonary hypertension is not reversed in an important subset of patients with severe mitral regurgitation after mitral valve replacement surgery and it is associated with a poor prognosis. No prospective study has been published on preoperative factors associated with this condition. The prevalence and prognosis of postoperative PH has not been precisely described.

*What this study adds:* Persistent PH after valve replacement for severe mitral regurgitation is common and its prognosis is ominous. Preoperative SPAP ≥50 mmHg and persistent atrial fibrillation are the main predictive factors. The results of this study might affect clinical practice suggesting considering surgery in this population (not only in mitral valve repair) when SPAP ≥50 mmHg, especially if persistent atrial fibrillation.

**Graphical abstract:** 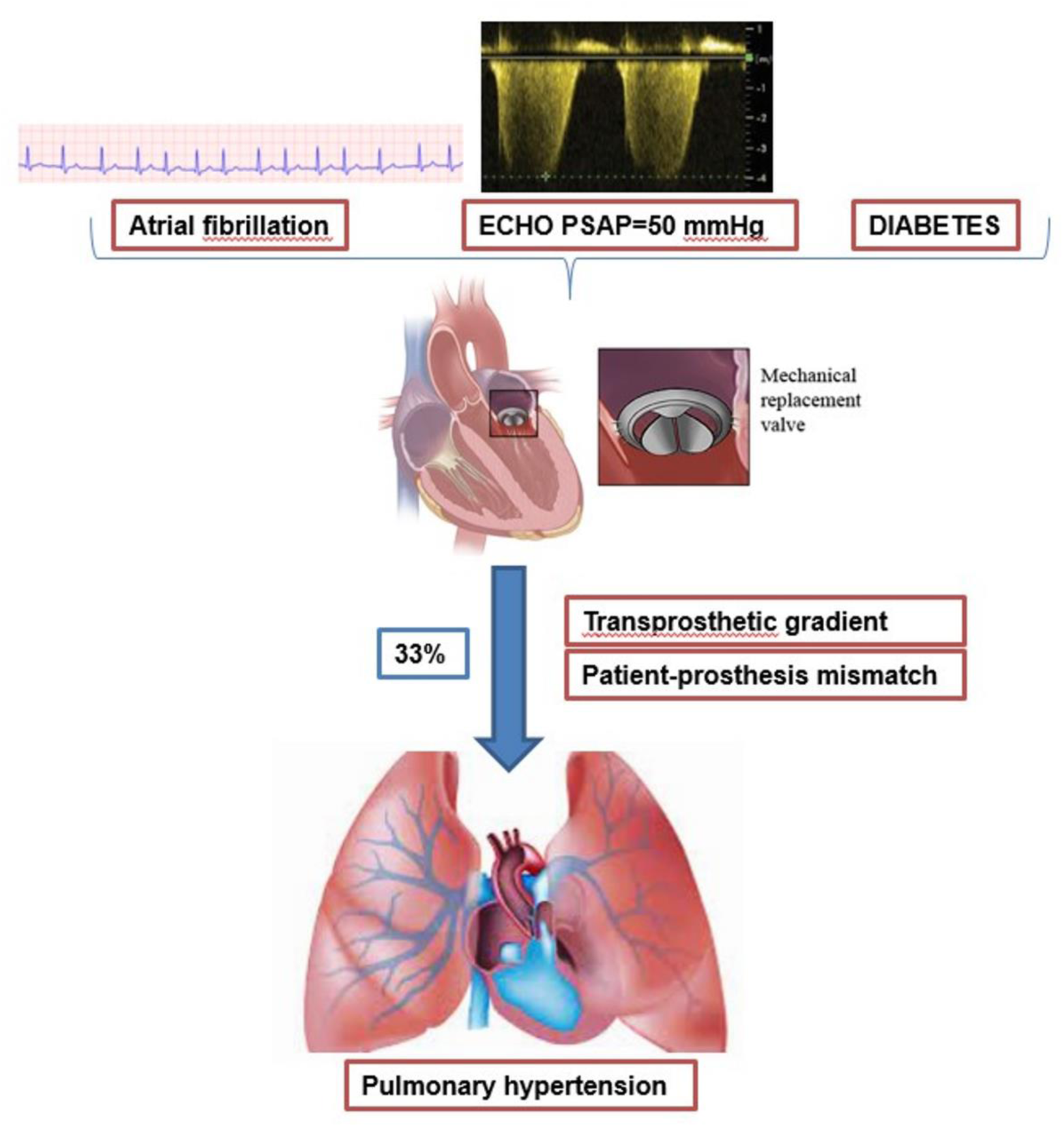

## INTRODUCTION

Mitral regurgitation (MR) is the second most frequent heart valve disease in Europe. Surgical repair is the gold standard approach to primary valve lesions but is not performed in half of the patients undergoing surgery for MR in Europe and North America, who are treated with prosthetic replacement (1–3).

Mitral valve disease is associated with pulmonary hypertension (PH), which is in principle secondary to passive retrograde transmission of increased left atrial pressure to the pulmonary capillary system (post-capillary PH). Successful surgical treatment of mitral valve disease is usually associated with the resolution of PH, whose persistence can reduce the exercise capacity of patients and increase their morbidity and mortality (4,5). In a subset of patients, however, PH does not disappear after successful valve surgery and can even worsen (6,7). There has been little in-depth study of perioperative factors that predict PH persistence after mitral valve replacement.

The main objective of this study was to determine the preoperative clinical and echocardiographic factors associated with PH persistence after mitral valve replacement. A secondary objective was to evaluate the effect of postoperative PH on the long-term prognosis.

## STUDY DESIGN AND METHODS

### 1. Design and sample

Patients undergoing valve replacement were consecutively recruited between January 2005 and January 2015 from a prospective cohort of patients with severe organic MR undergoing surgery at a referral center for a population of 1,620,000 inhabitants. Selected patients were followed up prospectively according to a pre-established protocol with preoperative, postoperative, and annual clinical and echocardiographic examinations.

### 2. Inclusion/exclusion criteria

The inclusion criterion was the receipt of replacement surgery for severe organic MR during the study period. Exclusion criteria were the receipt of mitral repair surgery, age < 18 or > 80 years at surgery, the presence of moderate or severe mitral stenosis (mitral valve area ≤1.5 cm*²* by echocardiography), mitral prosthesis dysfunction (thrombosis or prosthetic or periprosthetic regurgitation), and absence of postoperative tricuspid regurgitation for systolic pulmonary artery pressure (SPAP) estimation by echocardiography.

### 3. Variables

Data gathered on clinical and echocardiographic characteristics of patients are reported in supplemental material (e-table 1).

**Table 1:** Baseline characteristics of patients.

| Variables | n (%) |
| --- | --- |
| **Baseline characteristics** |  |
| Age (mean; sd) | 61.75; 11.15 |
| Sex (male) | 49 (46.2%) |
| Hypertension | 34 (32.1%) |
| Diabetes | 13 (12.4%) |
| Kidney failure | 3 (4.8%) |
| COPD | 8 (7.5%) |
| Smoking habit | 14 (14.1%) |
| **Cardiological history** |  |
| NYHA>2 | 71 (68.3%) |
| Admissions with heart failure | 31 (52.5%) |
| Preoperative atrial fibrillation | 64 (60.4%) |
| Coronary disease | 11 (10.4%) |
| Previous cardiac surgery | 14 (13.5%) |
| EuroSCORE Me[P<sub>25</sub>-P<sub>75</sub>] | 5 [3-6] |

COPD: Chronic obstructive pulmonary disease. NYHA: New York Heart Association.

### 4. Data gathering

A study protocol was followed to gather data on the clinical characteristics of patients, preoperative echocardiogram results, surgical variables, perioperative complications, and results of clinical and echocardiographic examinations at one-month post-surgery and then annually. Baseline and follow-up echocardiographic studies were conducted by two expert echocardiographers.

- <u>Independent data access and analysis</u>: the corresponding author had full access to all the data in the study and takes responsibility for its integrity and the data analysis.

### 5. SPAP estimation and definition of persistent PH

Echocardiography was used to estimate SPAP in all patients and an additional hemodynamic study in a subset of patients. Persistent PH was considered when pulmonary pressure by echocardiography was >50 mmHg (moderate PH = >50 mmHg, severe PH = >60 mmHg) at least 12 months’ post-surgery. Right atrial pressure was derived from the diameter and respiratory changes of the vena cava.

A hemodynamic study was performed in 20% of patients. Diastolic pressure gradient and transpulmonary pressure gradient were calculated by subtracting the mean pulmonary artery wedge pressure (PAWP) from the mean diastolic and pulmonary artery pressures, respectively. Pulmonary vascular resistance (PVR) and pulmonary arterial compliance were calculated as transpulmonary pressure gradient/cardiac output and stroke volume/ pulse pressure, respectively. PH was defined by mPAP ≥20 mm Hg and classified according to hemodynamic criteria as precapillary PH (PAWP ≤15 mm Hg,) isolated postcapillary PH (PAWP >15 mm Hg and PVR <3 Wood units [WU]) or combined precapillary and postcapillary PH (PAWP >15 mm Hg and PVR ≥3 WU).

A systematic review and meta-analysis comparing diagnostic accuracy between echocardiography and right cardiac catheterization reported a sensitivity of 88%, specificity of 90% and area under the ROC curve of 0.94 for echocardiography (4).

### 6. EOA estimation

The effective mitral valve area (EOA) was determined by the continuity equation. Patient-prosthesis mismatch (MM) was defined by an effective valve area ≤1.2 cm^2^/m^2^ of body surface area.

### 7. Statistical analysis

In relation to the main study objective, frequencies were calculated for categorical variables, means and standard deviations for quantitative variables with normal distribution, and medians and interquartile ranges for those with non-normal distribution (Kruskal Wallis test). Bivariate analyses were conducted to study the association between the response variable and each covariable or predictor variable. Given that the response was dichotomous (presence/absence of PH), the association of response with categorical predictors was assessed by estimating the relative risk, while the association with quantitative predictors was evaluated by estimating the odds ratio with a simple binary logistic regression model. A multivariate binary logistic regression model was constructed to evaluate the combined effect of predictors. Given the large number of predictors, only clinical variables significantly associated with the response in bivariate analysis were entered in the model. The discriminative capacity of the model was measured by calculating the area under the receiver operating characteristic (ROC) curve.

Regarding the secondary objective, mortality, readmissions, functional degree, and complications were compared between patients with *versus* without PH. The Kaplan-Meier procedure was used to estimate the survival of all patients, and survival was compared between those with *versus* without PH using the log-rank test.

R 4.0.1 software with survival package was used for data analyses, setting the level of significance at 5%.

### 8. Ethical aspects

The study complied with the Declaration of Helsinki for human investigation and was approved by the City Ethics Committee. All patients signed informed consent to their participation in the study. The corresponding author had full access to all the data in the study and take responsibility for its integrity and the data analysis.

## RESULTS

The study included 106 patients enrolled at hospital discharge, representing 87% of all patients undergoing valve replacement during the study period (127p). The causes of non-inclusion were associated mitral stenosis in 8.2% (10p), impossibility to estimate PASP in 4.9% (6p), and age >80 years in 4% (5p).

### 1. Patients characteristics

Table 1 lists the baseline characteristics and cardiac history of patients. Hypertension was observed in 32.1%, previous admission for heart failure in 52.5%, and NYHA functional class III or IV in 68.3% of patients; 60.4% were in permanent AF at enrolment.

Rheumatic etiology was the most frequent (50% of patients), followed by degenerative etiology (27.5%), and endocarditis (10%). Mitral valve surgery was performed alone in 47.1% of patients, with aortic surgery in 23.11%, and with tricuspid surgery in 22.6%. Revascularization surgery was carried out in 1.9%.

Follow-up ranged between 3 and 16 years, with a mean of 8.8 years.

### 2. Postoperative PH

PH appeared during the follow-up in 33 patients with pulmonary pressure >50 mmHg confirmed by echocardiography. Baseline SPAP was >60 mmHg in 17% of patients. Transesophageal echocardiography was routinely performed in patients with suspicion of prosthetic dysfunction, based on clinical or transthoracic echocardiography findings, and in all patients with PH >50 mmHg by echocardiography.

PH was confirmed in the subgroup of patients (20% of total, the first 25 patients included) who underwent hemodynamic study (mPAP ≥ 20 mmHg in all cases). Mean pulmonary pressure was 38 mmHg (33–47 mm Hg), considerably higher than the cutoff of 20 mmHg currently established as PH diagnostic criterion (8), mean pulmonary artery wedge pressure was 24 mmHg (19–27 mmHg), and median PVR was 3.3 WU (2.2–4.8 WU). Precapillary PH was detected in 5%, postcapillary PH in 40%, and combined precapillary/postcapillary PH in 55%.

### 3. Clinical and echocardiographic predictors of pulmonary hypertension

e-table 2 exhibits the results of bivariate analyses of the relationship between clinical or echocardiographic variables and PH persistence. Among preoperative variables, a statistically significant association was obtained with age, arterial hypertension, diabetes, or SPAP>45 mmHg. Associations with NYHA functional class>2, previous admissions for heart failure, and persistent AF were close to statistical significance. Among postoperative echocardiographic variables, a significant association was found with mitral prosthetic gradient. No significant between-group differences were observed in the type or number of prostheses or in the effective area of the prosthesis, either indexed or not.

**Table 2.** Multivariate logistic regression analysis.

|  | **OR** | **CI (95%)** |  | **p** |
| --- | --- | --- | --- | --- |
| (Constant) |  | 0.000 | 0.004 | 0.001 |
| Diabetes | 8.382 | 0.950 | 73.970 | 0.056 |
| Persistent AF | 4.250 | 0.876 | 20.618 | 0.053 |
| MR etiology | 7.378 | 1.196 | 45.529 | 0.031 |
| SPAP ≥50mmHg | 19.335 | 3.475 | 107.584 | 0.001 |
| Mismatch | 16.970 | 1.229 | 234.237 | 0.034 |
| Final gradient | 2.840 | 1.399 | 5.767 | 0.004 |

AF: atrial fibrillation MR: Mitral regurgitation. SPAP: systolic pulmonary artery pressure.

Postoperative PH was not associated with the area of the mitral prosthesis estimated by continuity equation, either absolute or relative to the body surface.

Variables showing statistical significance in bivariate analysis and others deemed potentially relevant were entered in the multivariate model, which identified the following predictive factors: diabetes, persistent AF, non-rheumatic MR, transprosthetic gradient, patient-prosthesis MM, and preoperative SPAP ≥50 mmHg. The final model is displayed in Table 2. Table 3 exhibit the ranges of measurements obtained for prosthetic area measurement and mistmatch in the whole series and in the groups with and without PH. Similar relationships are shown in e-tables 3 and 4 for other preoperative and predischarge echocardiographic variables, and associated lesions.

**Table 3.** Prosthetic area measurement (cm) and mismatch in the population and in patients with or without pulmonary hypertension.

|  | MEAN | STD. DEV | MIN | MAX | RANGE |
| --- | --- | --- | --- | --- | --- |
| **PROSTHESIS AREA** |  |  |  |  |  |
| PH absent | 2.024 | 0.25 | 1.6 | 2.7 | 1.1 |
| PH present | 2.1 | 0.38 | 1.3 | 2.8 | 1.5 |
| ALL | 2.0 | 0.3 | 1.3 | 2.8 | 1.5 |
| **PROSTHESIS MISTMACH** |  |  |  |  |  |
| PH absent | 1.27 | 0.24 | 0.84 | 2.0 | 1.16 |
| PH present | 1.31 | 0.31 | 0.74 | 1.96 | 1.22 |
| ALL | 1.28 | 0.27 | 0.74 | 2.0 | 1.26 |

**Table 4.** Mortality and complications as a function of postoperative PH.

|  | **PH present** | **PH absent** | **RR** | **p** |
| --- | --- | --- | --- | --- |
| Mortality | 24 (72.7%) | 13 (19.4%) | 2.96 (1.67-5.22) | 0.01 |
| Readmission | 26 (78.8%) | 25 (37.3%) | 2.96 (1.49-5.85) | 0.01 |
| NYHA >II | 26 (81.3%) | 41 (62.1%) | 2.02 (0.92-4.43) | 0.07 |
| Heart failure | 11 (68.8%) | 15 (40.5%) | 2.29 (0.92-5.67) | 0.08 |
| Complications | 8 (61.5%) | 8 (25%) | 2.90 (1.18-7.39) | 0.02 |

Complications: Heart failure, pericardial effusion requiring intervention, endocarditis, prosthetic dysfunction, sternal dehiscence requiring intervention, stroke, dysfunction of another prosthesis, fever requiring admission.

The area under the ROC curve (discriminatory capacity) of the final model was 0.873, as depicted in Figure 1.

**Figure 1.**
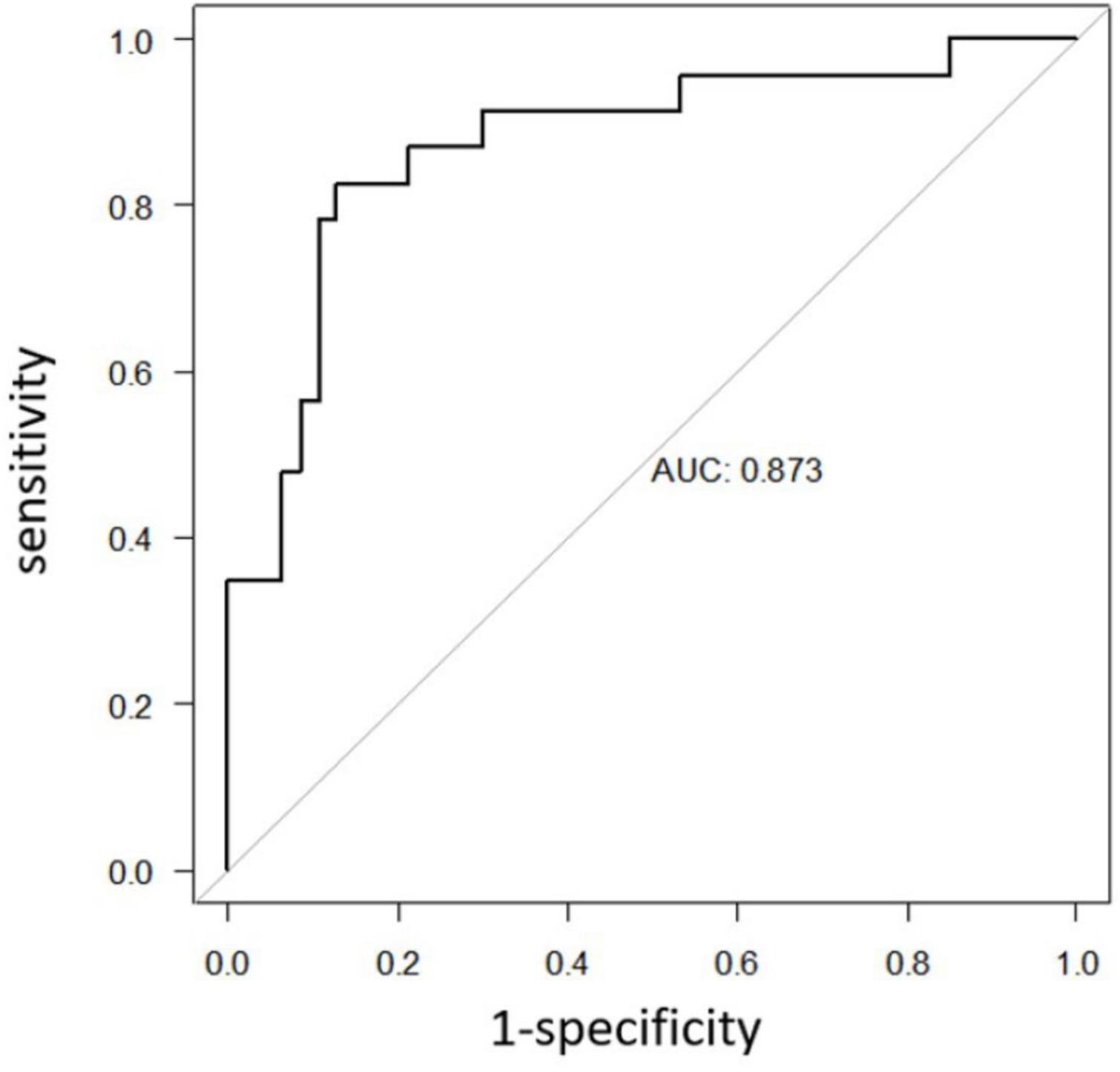
ROC curve. The multivariate model identified diabetes, persistent AF, non-rheumatic MR, transprosthetic gradient, patient-prosthesis MM, and preoperative SPAP ≥50 mmHg as predictive factors. The area under the ROC curve (discriminatory capacity) of the final model was 0.873. AF: atrial fibrillation. MM: mismatch. MR: mitral regurgitation. SPAP: systolic pulmonary artery pressure. ROC: receiver operating characteristic.

### 4. Postoperative PH and prognosis

With respect to the secondary objective of the study, patients with PH persistence had a higher frequency of readmissions (78.8 vs. 37.3%), poor functional class (NYHA III-IV 81.3 vs. 62.1%), admissions in heart failure (68.8 vs. 40.5%), and complications (61.5 vs. 25 %), and they had a markedly higher mortality rate (72.7 vs. 19.4%), as shown in Table 4.

### 5. Survival analysis

The survival analysis evidenced a statistically significant difference between patients with *versus* without PH (p<0.001) as displayed in figure 2.

**Figure 2:**
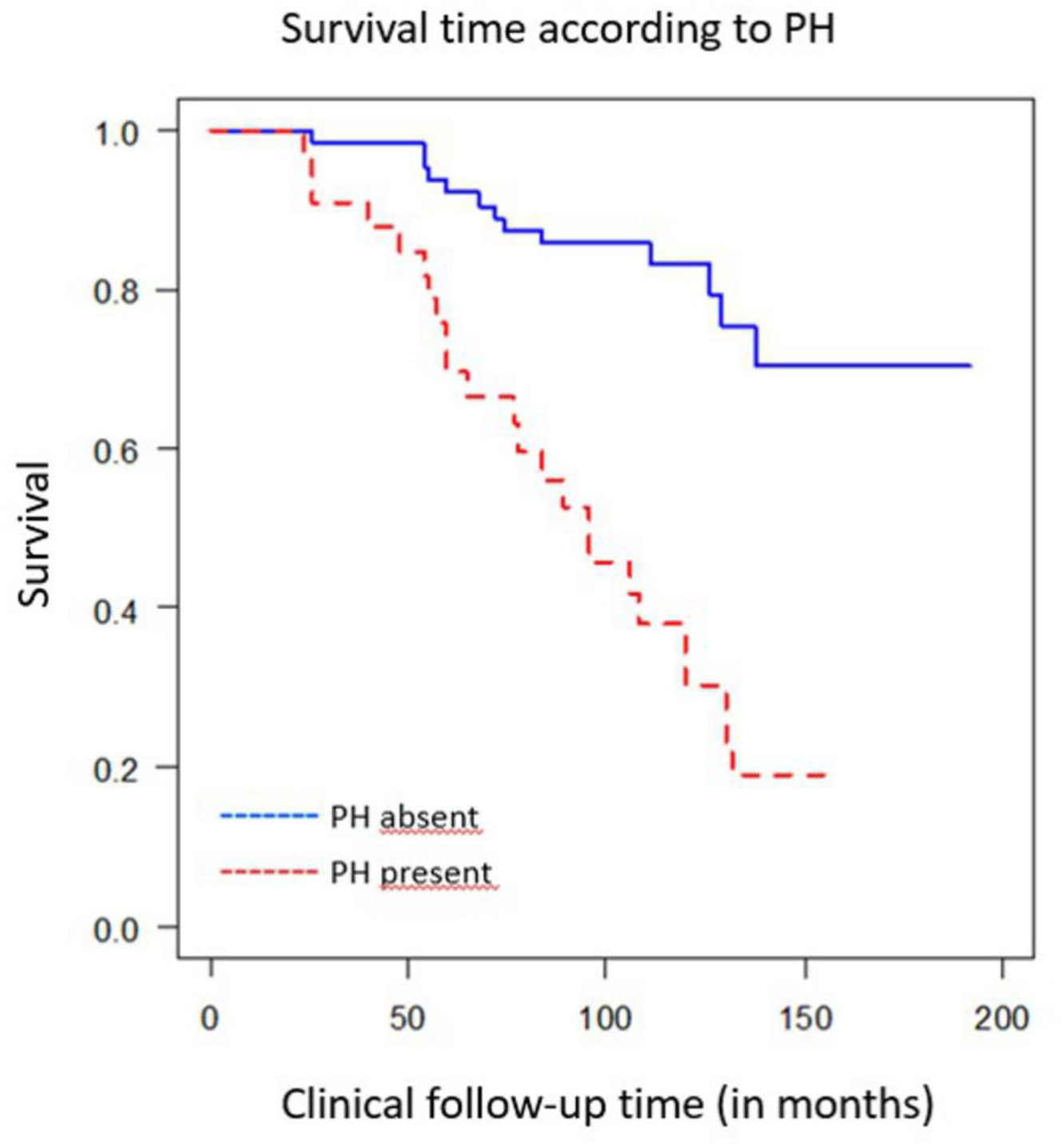
Survival as a function of the presence or absence of PH. The survival analysis evidenced a statistically significant difference between patients with versus without PH. Figure 2 depicts the survival according to the presence (dotted line, red) or absence (blue line) of PH. Survival decreases dramatically from the first months and throughout the follow-up. PH: pulmonary hypertension.

Tricuspid regurgitation was not significantly associated with mortality, unlike previous reports by some authors (9,10).

A Cox multivariate model was constructed for mortality-related parameters during the follow-up, entering variables that were significant in the bivariate analysis or considered relevant. Three predictive factors emerged for mortality: age, persistent AF, and preoperative PH (table 5). The mortality risk was four-fold higher in patients with *versus* without PH.

**Table 5.**
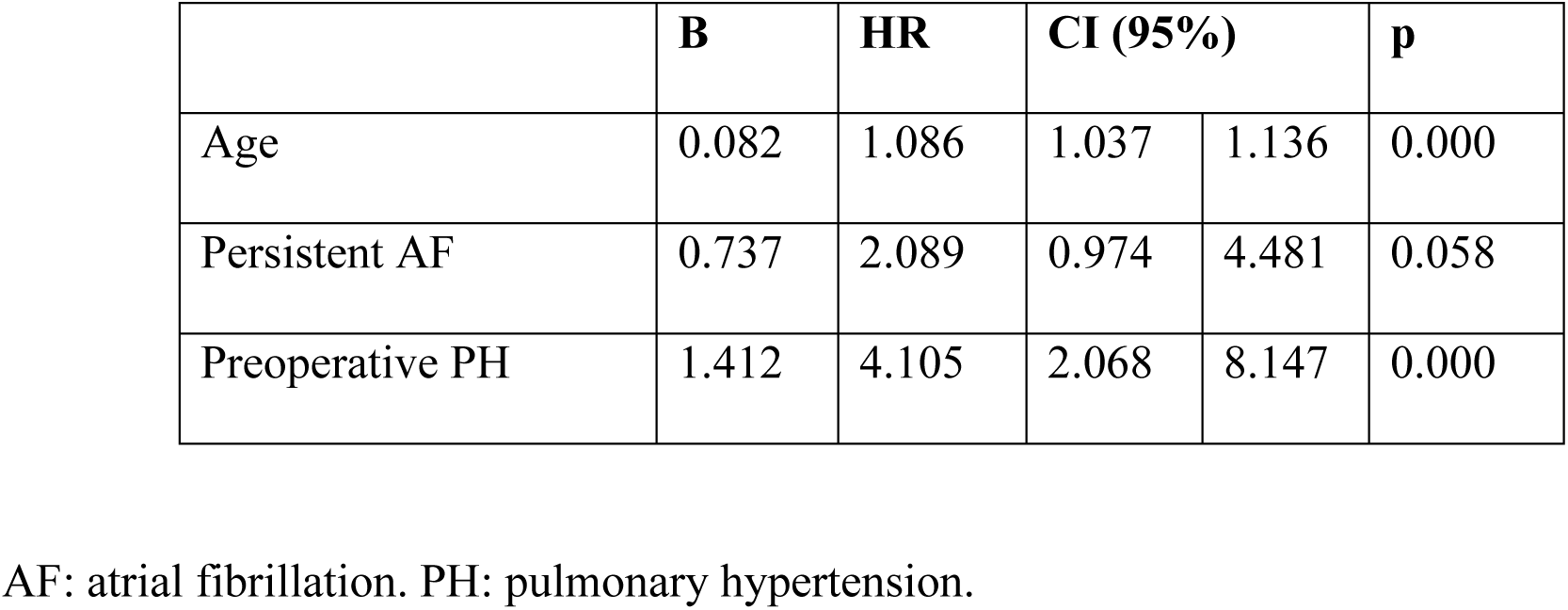
Mortality-associated factors during follow-up.

## DISCUSSION

Although mitral valve repair is the surgery of choice to treat severe mitral regurgitation, a substantial number of patients in the Western world continue to undergo valve replacement with a prosthesis, as confirmed by a recent survey (3).

This study had a long-time inclusion period because it selected all patients not candidates for mitral repair surgery, and it covered a long follow-up to detect PH. This may explain why the predominant etiology was rheumatic valve disease. However, 27.5% of the patients with degenerative regurgitation also underwent valve replacement. These patients frequently have multivalvular disease or other associated surgery, as in the present series.

Persistent PH after successful mitral valve replacement changes the pulmonary vascular bed and is associated with increased post-operative morbidity and mortality (5,7). To our best knowledge, no prospective study has been published on perioperative factors associated with the onset or persistence of PH. These data could support decision-making on the indication of surgery in patients undergoing mitral valve replacement for which the published evidence is very scarce unlike that related to mitral repair surgery.

The main limitation of studies on this issue is their retrospective design, implicated in the failure to consider preoperative variables, the enrolment of followed-up rather than consecutive patients, the absence of systematic echocardiographic studies during follow-up, with consequent losses and biases, and follow-up periods that are too short to fully evaluate the presence of PH and the impact on outcomes. Furthermore, studies have frequently included patients with both regurgitation and an advanced degree of mitral stenosis, which may have distinct outcomes.

### 1. Prevalence of PH after mitral valve replacement

The prevalence of postoperative PH among published series is highly variable, ranging from 22 to 68% (11–13). The present study, with a prospective and rigorous annual clinical and echocardiographic follow-up, found a prevalence of 33% over a period of more than 10 years, associated with a markedly higher mortality rate and a much greater percentage of complications.

### 2. Factors associated with persistent PH

Various factors have been associated with persistent PH, although study findings have widely varied. Patient-prosthesis MM and the degree of pre-surgical PH have been implicated by some studies but not others, as is the case for the transprosthetic gradient. Other proposed predictive factors include severe preoperative PH (> 60 mmHg), mitral prosthesis area, atrial fibrillation, and pre- and post-operative left ventricular function parameters. The preoperative factors associated with postoperative PH persistence or progression in the present multivariate analysis were diabetes, persistent AF, non-rheumatic etiology, and preoperative PH ≥50 mmHg (by echocardiography), which was found to carry a 20-fold higher risk of postoperative PH in comparison to the absence of pre-operative PH. Among perioperative parameters, MM and transprosthetic mitral gradient were associated with postoperative PH. No association was found with other factors that have been related to this condition, such as the mitral prosthesis area indexed to body surface area, pre- or postoperative left ventricular function parameters, or a history of lung disease.

### 3. Patient-prosthesis mistmach

MM of the mitral prosthesis, defined as an indexed EOA ≤ 1.2 cm²/m², has been implicated in postoperative PH through elevation of the transprosthetic gradient, which would delay or prevent the resolution of atrial dysfunction and PH. However, this association was not observed in some other studies (11–20). A review of publications since 2018 retrieved 103 related articles, all retrospective. Among the 18 studies considered to offer high-quality evidence, eight reported a negative impact of MM on mortality whereas five found no association. Some authors observed an association between MM and higher postoperative pulmonary pressure. Other authors proposed that MM reflects a higher baseline risk but is not itself responsible for a poor prognosis. In the present study, which controlled for multiple potential confounders, a positive relationship was found between PH and moderate or severe MM over the long-term.

### 4. Severe preoperative PH

Previous severe PH (≥60 mmHg) may also be associated with postoperative PH persistence. However, various studies have shown that these patients may have greater perioperative mortality but do not have higher pulmonary pressure during their follow-up (24). This is supported by the present finding of no significant association between severe PH at baseline and PH persistence during the follow-up.

### 5. Atrial fibrillation

The onset of AF has been described as a predictor of cardiac morbidity and mortality in patients with degenerative MR undergoing surgery (25). In the present patient population, AF was highly prevalent and found to be related to postoperative PH and to predict a poor outcome.

### 6. Mitral transprosthetic gradient

Increased mitral transprosthetic gradient was significantly associated with postoperative PH in the present study, regardless of the EOA of the prosthesis, whether or not indexed to body surface area. Other authors found no correlation between transprosthetic gradient and prosthesis size. It could be hypothesized that this gradient is related to left atrial function and pressure, with subsequent passive transmission to the pulmonary tree. In this way, left atrial dysfunction would raise the intra-atrial pressure, which would increase venous congestion and therefore favor PH (25).

### 7. Guidelines recommendations

The above controversial evidence forms the basis of the most recent guidelines on surgical indications for valvular heart disease (26). In American guidelines (27), surgery is only indicated for asymptomatic patients with systolic pressure ≥50 mmHg if they do not have rheumatic valve disease and are candidates for repair surgery but not valve replacement; in the subgroup of candidates for valve replacement, patients must meet criteria for symptoms or for ventricular dysfunction, but the presence of isolated PH is not a criterion for surgery. In European guidelines (28), there is a general recommendation for surgery in asymptomatic patients with SPAP >50 mmHg, based on evidence from two studies by MIDA investigators; however, the MIDA registry included patients with mitral regurgitation due to flail mitral valve leaflets, and 90% of these underwent mitral valve repair (29,30). Hence, this recommendation is supported for patients scheduled for valve repair but not for valve replacement candidates.

In the present study, there was a high prevalence of long-term PH after valve replacement, and preoperative SPAP ≥50 mmHg emerged as an independent predictor of this outcome. Morbidity and mortality rates were very high in affected patients. These findings suggest that surgery may be indicated in patients with moderate PH who are not candidates for valve repair to prevent postoperative PH and the associated morbidity and mortality, in the same way as recommended in candidates for mitral valve repair. It is not ethically feasible to conduct a randomized study to test this hypothesis; however, these findings should serve to alert clinicians to the poor prognosis of these patients after replacement, and to intensify the search for symptoms and any indications for surgery in patients with moderate PH. Surgery would be especially indicated in patients with degenerative MR and those with permanent AF.

The presence of MM and augmented gradient after prosthesis implantation are also predictors of PH during follow-up and therefore of a poor prognosis.

### 8. Limitations

The relatively small sample size reduced the statistical power of this study, and some of the associations considered may reach statistical significance in larger samples.

These patients underwent mitral valve replacement with mechanical prosthesis, and the results cannot be extrapolated to patients undergoing mitral valve repair surgery or percutaneous treatment.

The predictive capacity of the model was evaluated using the same individuals who served for its construction rather than a validation cohort. This implies a possible overestimation, although this does not usually exceed 10-15% and the present results would therefore remain significant. The number of patients was not sufficient to provide a test set for the assessment of factors in the multivariate analysis.

Hemodynamic study is the gold standard diagnostic test for PH. It was not conducted in all patients because the study objective was not to test the diagnostic accuracy of transthoracic echocardiograms for PH but rather to identify predictors of postoperative PH in relation to the surgical indications in clinical practice guidelines, none of which indicate hemodynamic measurements. Nevertheless, the first 25 patients enrolled in the study underwent hemodynamic study to evaluate the accuracy of echocardiographic estimation in this specific subgroup, which was verified in all 25 patients, with results clearly above the cutoff point.

### Conclusions

Persistent PH after mitral valve replacement is associated with the presence of moderate preoperative PH. Long-term mortality and morbidity rates are very high in patients with postoperative PH. These findings suggests that valve replacement surgery may be considered in MR patients with moderate PH who are not candidates for valve repair, especially in the presence of atrial fibrillation.

## Data Availability

All data generated or analyzed during this study are included in this published article and its supplementary information files. Additional data that support the findings of this study are available from the corresponding author upon reasonable request.

## ABBREVIATIONS

HP: Pulmonary hypertension
MR: Mitral regurgitation
MM: Patient-prosthesis mismatch
SPAP: Systolic pulmonary artery pressure
EOA: Effective mitral valve area
AF: Atrial fibrillation.

## Acknowledgments, sources of founding and disclosures

**Guarantor:** RGO takes responsibility for the content of the manuscript, including the data and analysis

**Author contributions:** RGO and EME: Had access to all the data. They take responsibility por the integritiy of the data and the accuracy of the data analysis. In charge of the conceptualization, data analysis and original draft preparation: MCG, IUM, LTQ, DJRT AND DSR: Data gathering, data interpretation, draft review & editing.

**Sources of founding:** The corresponding author, on behalf of all the authors of a submission declares that this research received no external financial or non-financial support

**Disclosures:** There are no additional relationships to disclose. There are no patents to disclose. There are no additional activities to disclose

## Central figure-Graphical abstract

Persistent PH is a frequent long-term complication with a poor prognosis PH after mitral valve replacement. It appeared in 33% of patients after a median follow up of 8.8 years. Predictor factors were diabetes, persistent atrial fibrillation, transprosthetic gradient, patient-prosthesis mismatch and preoperative SPAP≥50 mmHg.

